# The Mental Health of Healthcare Staff during the COVID-19 Pandemic: It Depends on How Much They Work and Their Age

**DOI:** 10.1101/2020.08.18.20173500

**Authors:** Xingzi Xu, Stephen X. Zhang, Asghar Afshar Jahanshahi, Jizhen Li, Afsaneh Bagheri, Khaled Nawaser

## Abstract

**Background:** Healthcare staff are the forefront of fight against COVID-19 and they are under enormous pressure due to the fast growth in the number and severity of infected cases. This creates their mental issues such as distress, depression and anxiety. Exploring healthcare staff’s mental health during the pandemic contributes to improving their persistence in the growing challenges created by COVID-19 and enabling effective management of their mental health.

**Methods:** An online survey of 280 healthcare staff in all the 31 provinces of Iran was conducted during April 5–20, 2020. The survey assessed staff’s distress, depression and anxiety in the COVID-19 pandemic.

**Results:** Nearly a third of healthcare staff suffered from distress, depression and anxiety. Females and more educated healthcare staff were more likely to experience distress. Compared to personnel who did not have COVID-19, those who were unsure whether they had COVID-19 were more likely to experience distress and depression. The number of COVID-19 cases among the staff’s colleagues or friends positively predicted their anxiety. Compared to radio technologists, doctors were less likely to experience distress and anxiety. Technicians and obstetrics experienced less anxiety. Analysis the interaction between weekly working days and age of the staff indicated the chance of experiencing distress and depression varied greatly by working days among younger but not older healthcare staff.

**Conclusion:** The predictors of mental health issues assists healthcare organizations to identify healthcare staff with mental health issues in sever crises such as the COVID-19 pandemic. Our research highlight the need to identify more working characteristics as predictors for healthcare staff at different ages.

**Funding:** This work was supported by Tsinghua University-INDITEX Sustainable Development Fund (No. TISD201904).

## 1. Introduction

Healthcare staff have been under enormous physical and mental pressures since the beginning of the COVID-19 pandemic. In the pandemic, healthcare staff face tremendous workloads, long and irregular working hours, and shortages of personal protective equipment (PPE) (Woolley, Smith, & Arumugam, 2020). They also experience emotional and moral dilemmas from being isolated from their families during the pandemic (Lee, 2020; Roy et al., 2020). All of these factors may eventually lead to mental health issues (Afshar et al., 2020; Lai et al., 2020). For instance, one nurse committed suicide because of the overload stress in Italy (Ruiz, 2020). “Staff are being exposed to high levels of personal risk, long hours in difficult environments clad in PPE, and also the possibility of something known as moral injury, which is the distressing awareness you may feel when you know you can’t meet all the needs of the people you are trying to care for” (Jane, 2020). While such issues are acute in many countries (Dai, et al., 2020; Zhang, Sun, Jahanshahi, Alvarez-Risco, et al., 2020), this paper aims to specify some unique predictors of healthcare staff’s mental health disorders in Iran as the second country that experienced COVID-19 and the virus spread in all of the country very fast.

In predicting mental health issues created and/or developed by COVID-19, this current research focused on socio-demographic (Byrne, 2020; Henry & Lippi, 2020) and clinical predictors of healthcare staffs (Banerjee,2020). This survey is one of the very first attempts that investigated working characteristic predictors of healthcare staff’s mental health during the COVID-19 pandemic. Furthermore, this study is the first, to the best of our knowledge, to examine comprehensive predictors of healthcare staff’s mental health by measuring socio-demographic, clinical and working variables.

While previous studies have suggested the risk factors as the predictors of mental health among healthcare staff (Chen J. et al., 2020; Chen X. et al., 2020; Zhang, Graf-Vlachy, Su, et al., 2020), our study particularly investigated working characteristics (Zhang, Wang, Rauch, et al., 2020) such as healthcare staff’s job function and their number of working days per week to predict their distress, depression and anxiety. We also tested how the interaction between weekly working days and age of the staff predicts these mental health issues. These unique predictors provide evidence to help psychological service to identify healthcare staff who may need mental health services.

## 2. Methods

Our study surveyed healthcare staff in Iran during April 5–20, 2020 when Iran was experiencing a high outbreak of COVID-19. On April 5, the date the survey started, a total of 58,226 cumulative COVID-19 cases were diagnosed in Iran and 2,483 were diagnosed as new cases. Of the diagnosed cases, 3,603 total deaths reported in a day, 43 of whom were healthcare staff (Gharebaghi & Fatemeh, 2020). On April 20, the date the survey ended, a total of 83,505 cumulative COVID-19 cases were reported in Iran with 1,294 daily new cases and 5,209 total deaths. This caused extreme strain in medical resources and great psychological pressures on Iranian healthcare staff.

### 2.1 Study Design

This study followed the American Association for Public Opinion Research (AAPOR) reporting guideline. Written informed approval was provided by all survey participants prior to their enrolment. In order to access more participants under the constraints created by the COVID-19 pandemic, we shared the survey links on the social media channels (Instagram, Whatsapp and Telegram) which are very popular among Iranian and asked the responding healthcare staff to share the survey with their colleagues. All of the participants completed the survey voluntarily and anonymously, and they were free to leave the survey at any time. The final sample comprised 280 healthcare staff across all the 31 provinces of Iran.

### 2.2 Measures

#### 2.2.1 Predictor variables

Our predictor variables included socio-demographic variables, clinical variables and working characteristic variables. Socio-demographic characteristics were self-reported by all participants including *age* (that ranged from 20–30, 30–40, 40–50, to 50–60 years old), *gender* (male or female), *education level* (categorical variable ranging from 1 = under diploma, 2 = diploma with 12 years of education, 3 = student or graduate with 2 academic years, 4=student or graduate from university, 5 = student or graduate with a master degree, 6 = student or graduate with a doctoral degree).

Clinical variables included: *chronic disease*, *infected situation* and *number of colleagues or friends diagnosed with COVID-19*. Drawing on previous research finding that people who have comorbidity may experience more mental health disorder than others in the COVID-19 pandemic (Fawad et al., 2020), we asked the participants in this survey whether they had chronic disease using a categorical variable (1=yes, 2=not sure, 3=no). The participants were also asked whether they had been infected with COVID-19 within the past two weeks (1=yes, 2=not sure, 3=no). Lastly, the participants reported the number of their colleagues or friends diagnosed with COVID-19 within the past two weeks.

Working characteristic variables consisted of three items. Because medical protective equipment are different by the job function (Rimmer, 2020), we asked the healthcare staffs to indicate their *job function* (a categorical variable ranging from 1 to 9, where 1= a doctor, 2= a nurse, 3= a technician, 4= a radio technologist, 5= a medical student and an intern, 6= a healthcare administrator, 7= a supporting staff such as facility or cleaning staff, 8= a volunteer and 9= a obstetric staff). We also asked the participants their *working institute* (private, public, or government sector) and how many days a week they worked as a proxy for *working days* (1 day to 5 days) because the variables were suggested to affect healthcare workers’ mental health.

#### 2.2.2 Outcome variables

The three outcome variables measured in this study were distress, depression, and anxiety. The Kessler Psychological Distress Scale (K6) with Cronbach’s alpha of 0.79 (Kessler et al., 2020) was used to assess the participants’ distress during the COVID-19. The 4-item Patient Health Questionnaire (PHQ-4, with a score of 0–12) was also employed to measure the severity of depression and anxiety among the participants. PHQ-4 is an ultra-brief self-report questionnaire consisting of a 2-item depression scale (PHQ-2) and a 2-item anxiety scale (GAD-2). The cutoff scores for detecting symptoms of distress, depression and anxiety were 13, 3 and 3 respectively. Participants who scored higher than the cutoff threshold were characterized as having the symptoms of the diseases. We translated the measures from English into Farsi, the official language of Iran.

### 2.3 Statistical Analysis

Our empirical analysis was performed using Stata 16.1 software. The statistical significance level was assessed by *p* < .05, 2-tailed. Multivariable logistic regression was used to determine potential risk threats of distress, depression and anxiety. In the statistical regression results, we presented descriptive analysis with odds ratios (ORs) and 95% confidence intervals (CIs) to show the potential association between risk predictors and outcome variables. We also performed a margin analysis to further investigate the interaction between the participants’ working days and age that predict their distress, depression and anxiety.

## 3. Results

### 3.1 Descriptive statistics

Table 1 displays the descriptive characteristics of the 280 healthcare staff participated in this survey. Of the staff, 21.4% (60) had distress disorder, 30.0% (84) had depression disorder, and 32.9% (92) had anxiety disorder. The majority of the participants (60.0%, 168) were female. Most of the participants (70.5%, 196) were under 40 years old, and 79.4% (239) of the participants had a university degree or 2-year diploma. Most of the participants (80.3%, 226) did not have chronic diseases, 2.5% (7) reported they were diagnosed as COVID-19 positive, while 27.1% (76) reported that they were unsure if they were infected by the virus. Over half of the participants (53.0%, 161) reported that their colleagues or friends were infected by COVID-19 with a mean of 3.86 (min: 0; max: 150). Because the number of colleagues or friends infected by the virus is a count, we transformed it by taking its log.

**Table 1.**
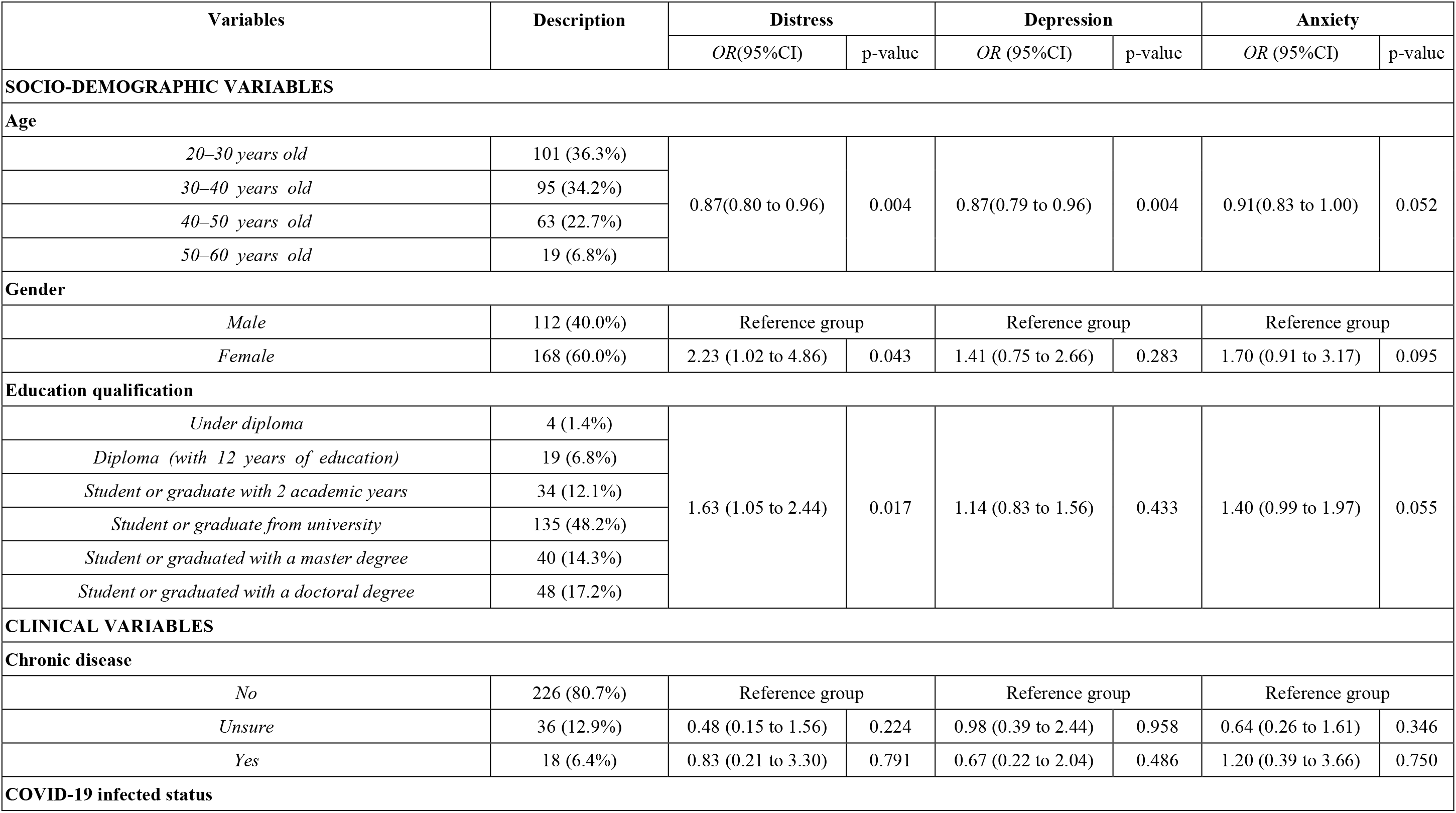
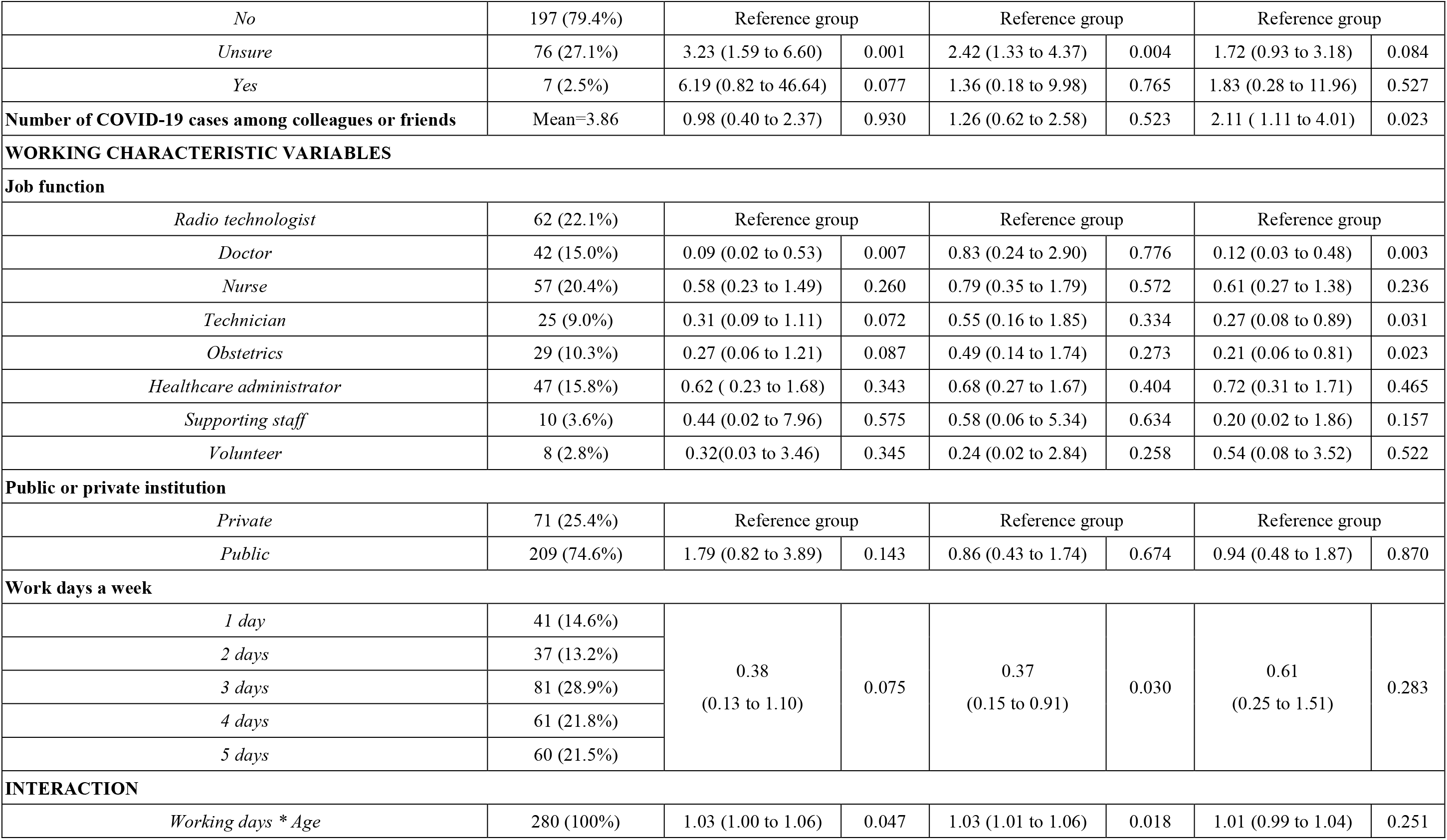
Descriptive findings and predictors of healthcare staff’s mental health by logistic regression (N=280)

Regarding the participants’ job functions, 15.0% (42) were doctors, 20.4% (57) were nurses, 9.0% (25) were technicians, 22.1% (62) were radio technologists, 3.6% (10) were medical students and interns, 13.2% (37) were healthcare administrators or interns, 3.6% (10) were supporting staff (i.e., facility or cleaning staff), 2.8% (8) were volunteers and 10.3% (29) were obstetrics staff. Most the healthcare staff were from a public healthcare organization (74.7%, 223) and worked at least three days a week (72.2%, 202).

### 3.2 Logistic regression results

Table 1 presents the multivariate logistic regression results. Firstly, our analysis revealed that females were 2.23 times more likely to have distress disorder (OR=2.23; 95% CI: 1.02 to 4.86; *p*=0.043) than their male counterparts. Highly educated healthcare staff experienced a higher level distress disorder (OR=1.63; 95% CI: 1.05 to 2.44; p=0.017) than less-educated staff. Compared to healthcare staff who were not infected by COVID-19, those who were unsure if they were infected by the virus had a higher level of distress (OR=3.23; 95% CI: 1.59 to 6.60; p=0.001) and depression disorder (OR=2.42; 95% CI: 1.33 to 4.37; p=0.004). The number of COVID-19 positive cases among colleagues or friends also positively predicted anxiety disorder (OR=2.11; 95% CI: 1.11 to 4.01; p=0.023). Compared to radio technologists – the biggest job function category in this study, medical doctors were less likely to experience distress (OR=0.09; 95% CI: 0.02 to 0.53; p=0.007) and anxiety disorder (OR=0.12; 95% CI: 0.03 to 0.48; p=0.003) while technicians (OR=0.27; 95% CI: 0.08 to 0.89; p=0.031) and obstetrics staff (OR=0.21, 95% CI: 0.06 to 0.81; p=0.023) were less likely to experience anxiety disorder. Our analysis also showed that existing chronic health issues and the type of healthcare organization (private, public or government sector) did not significantly predict distress, depression and anxiety. Significant results are presented in figure 1.

**Figure 1.**
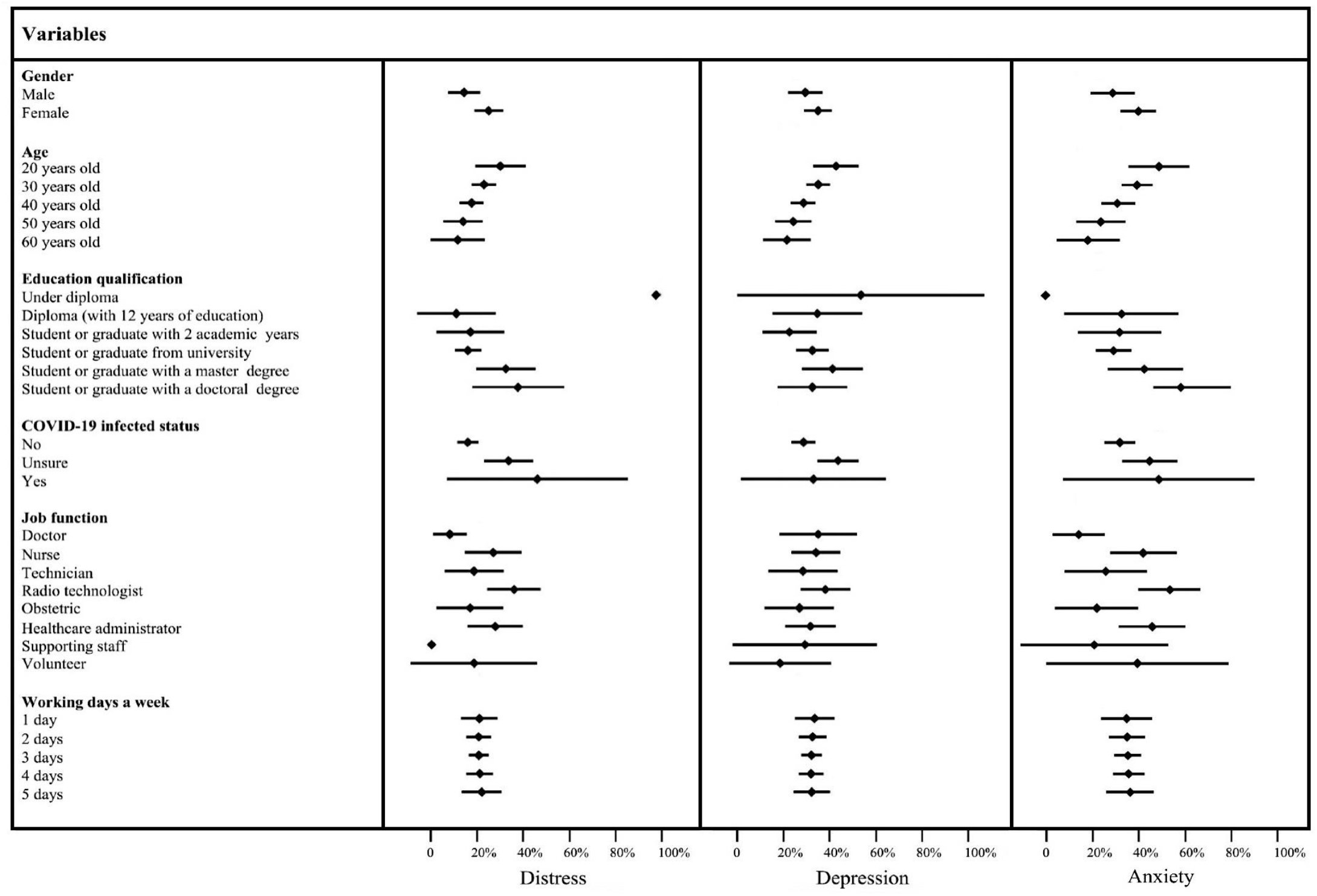
Predicted value and 95% confidence intervals (CIs) of distress, depression and anxiety by healthcare staff’s socio-demographic variables, clinical variables and working characteristic variables

Secondly, the results demonstrated that the interaction between the participants’ working days and age significantly predict their distress (OR=1.03; 95% CI: 1.00 to 1.06; *p*=0.047) and depression (OR=1.03, 95% CI: 1.00 to 1.06; *p*=0.026). Figure 1 shows the participants’ working days per week and age predict their distress and depression. By further conducting a margin analysis, we found that 41% of the healthcare staff who were 20 years old and worked only one day per week had the highest likelihood of distress (95% CI: 23% to 65%; *p*=0.000) and 61% of them experienced depression (95% CI: 40% to 82%; *p*=0.000). Furthermore, the healthcare staff who were 20 years old but worked five days a week had 19% (95% CI: 19% to 36%; *p*=0.023) chance of having distress disorder and 24% (95% CI: 7% to 40%; *p*=0.004) chance of depression disorder. The chance of having distress and depression varied significantly by the number of working days among younger participants but not their older counterparts. Margin analysis results are presented in figure 2.

**Figure 2.**
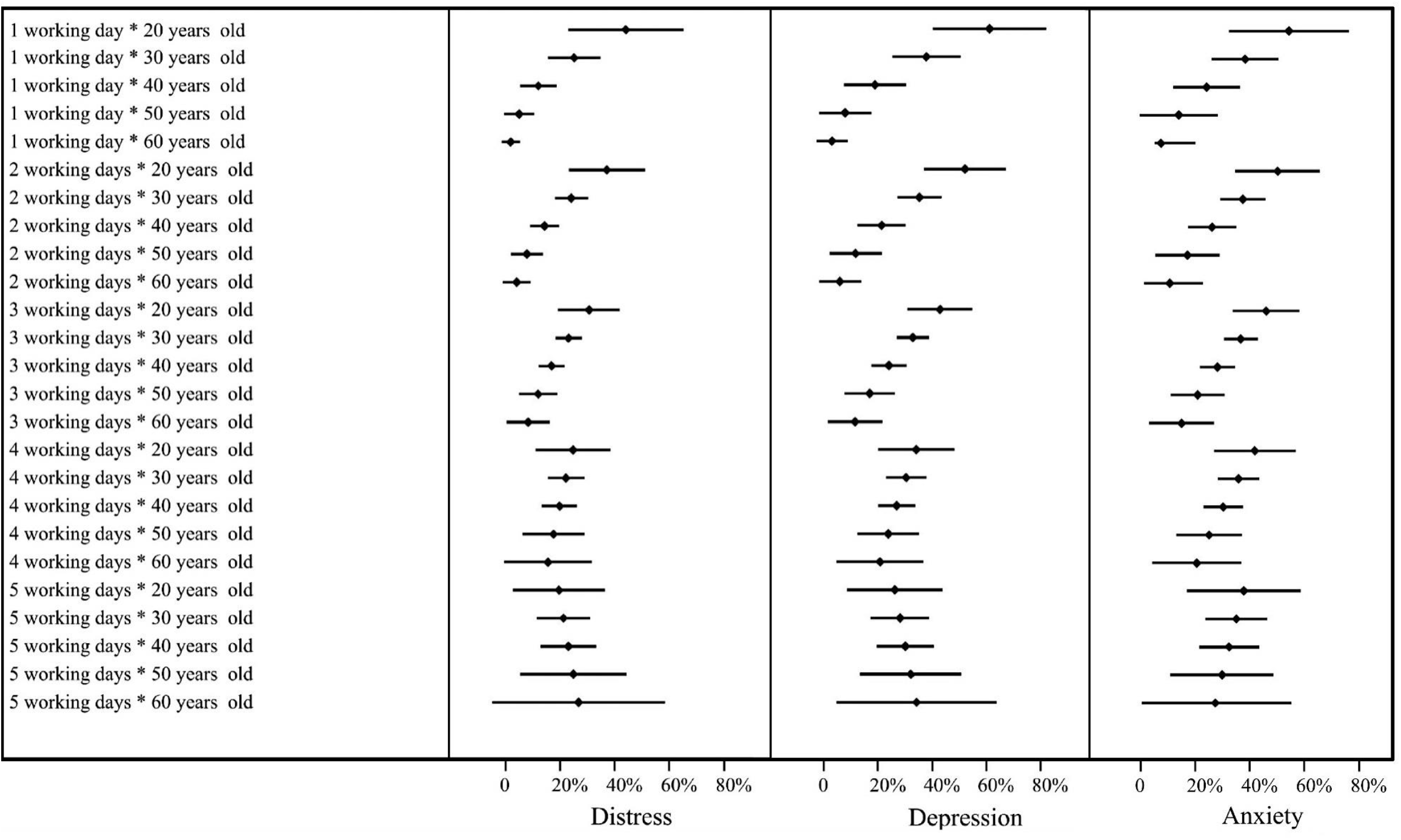
Predicted value and 95% confidence intervals (CIs) of distress, depression and anxiety by healthcare staff’s number of work days a week and age

## 4. Discussion

The findings of our study revealed that nearly a third of the healthcare staff in Iran reported symptoms of distress, depression and anxiety during the COVID-19 pandemic. Gender and education were significant predictors of the healthcare staffs’ distress. The participants who were unsure if they were infected by COVID-19 virus reported more distress and depression. In addition, our survey suggested that the number of COVID-19 cases among a healthcare staff’s colleagues or friends positively predicted his or her anxiety. Our study demonstrated that the interaction between the number of working days a week and age predicted healthcare staff’s distress and depression.

Some demographic predictors found by previous research failed to carry a significant effect in our study on the mental health of the healthcare staff. While, age, gender and education predicted the mental health among healthcare staff during the COVID-19 pandemic in China (Li et al., 2020; Xiao et al., 2020; Wang et al., 2020), Italy (Casagrande et al., 2020), Brazil (Zhang, Wang, Jahanshahi, et al., 2020) and several other countries (Zhang, Sun, Jahanshahi, Alvarez-Risco, et al., 2020), they were not significant in this study. In addition, previous studies indicated those who had a chronic disease had worse mental health (Sodhi & Manju, 2013; Hüfner et al., 2019), but such a relationship was not significant in this study. Our study also revealed that doctors were less likely to suffer from distress and anxiety than radio technologists. These findings may indicate that the predictors of the mental health of healthcare staff during the pandemic may vary across countries (Zhang, Liu, Jahanshahi, et al., 2020; Zhang, Sun, Jahanshahi, Wang, et al., 2020; Tang et al., 2020).

Interestingly, our study showed that the participants’ age predicted distress and depression differently depending on their working days per week. Our findings suggest younger healthcare staff who did not work much and older healthcare staff who worked a lot are the more mentally vulnerable groups. Unlike previous studies that have shown that younger adults experienced greater mood swings during the SARS outbreak (Yeung & Fung, 2020), we showed younger healthcare staff did not generally had worse mental health, and the prediction needs to take account of their working days. Our findings reveals how working days and age together jointly predicted the mental health of healthcare staff in the pandemic.

## 5. Limitations

Our study has several limitations that open new agendas for future studies. We attempted to include samples from different provinces of Iran. However, our sample is representative as we did not aim to capture healthcare staff by their proportions based on the provinces in Iran.

## 6. Conclusion

The predictors of mental health for healthcare staff assists psychological service institutions better identify the mentally venerable staff during the COVID-19 pandemic. The interaction of healthcare staff’s number of working days a week and age jointly predicted their distress and depression disorders. Our results suggest the need to further investigate healthcare staff’s working characteristics during the COVID-19 pandemic as predictors of their mental issues.

## Data Availability

we shared the survey links on the social media channels (Instagram, Whatsapp and Telegram) which are very popular among Iranian and asked the responding healthcare staff to share the survey with their colleagues.

